# Trajectories of genetic risk across dimensions of alcohol use behaviors

**DOI:** 10.1101/2025.03.27.25324798

**Authors:** Jeanne E Savage, Fazil Aliev, Peter B Barr, Maia Choi, Gabin Drouard, Megan E Cooke, Sally I Kuo, Mallory Stephenson, Sarah J Brislin, Zoe E Neale, Spit for Science Working Group, COGA Investigators, Antti Latvala, Richard J. Rose, Jaakko Kaprio, Danielle M Dick, Jacquelyn Meyers, Jessica E Salvatore, Danielle Posthuma

## Abstract

**Background:** Alcohol use behaviors (AUBs) manifest in a variety of normative and problematic ways across the life course, all of which are heritable. Twin studies show that genetic influences on AUBs change across development, but this is usually not considered in research identifying and investigating the genes linked to AUBs.

**Aims:** Understanding the dynamics of how genes shape AUBs could point to critical periods in which interventions may be most effective and provide insight into the mechanisms behind AUB-related genes. In this project, we investigate how genetic associations with AUBs unfold across development using longitudinal modelling of polygenic scores (PGSs).

**Design:** Using results from genome-wide association studies (GWASs), we created PGSs to index individual-level genetic risk for multiple AUB-related dimensions: *Consumption*, *Problems*, a variable pattern of drinking associated with a preference for beer (*BeerPref*), and externalizing behavior (*EXT*). We created latent growth curve models and tested PGSs as predictors of latent growth factors (intercept, slope, quadratic) underlying trajectories of AUBs.

**Setting:** PGSs were derived in six longitudinal epidemiological cohorts from the US, UK, and Finland.

**Participants:** Participant data were obtained from AddHealth, ALSPAC, COGA, FinnTwin12, the older Finnish Twin Cohort, and Spit for Science (total N = 19,194). These cohorts included individuals aged 14 to 67, with repeated measures collected over a span of 4 to 36 years.

**Measurements:** Primary measures included monthly frequency of typical alcohol consumption (CON) and heavy episodic drinking (HED).

**Findings:** Results indicated that higher PGSs for all AUBs are robustly associated with higher mean levels of CON and/or HED (B = 0.064-0.333, *p* < 3.09E-04). However, these same genetic indices were largely not associated with drinking trajectories across cohorts. In the meta-analysis, only PGSs for chronic alcohol *Problems* consistently predicted a steeper slope (increasing trajectory) of CON across time (B = 0.470, *p* = 4.20E-06).

**Conclusions:** The results indicate that genetic associations with AUBs not only differ between behaviors, but also across developmental time points and across cohorts. Genetic studies that take such heterogeneity into account are needed to better represent the underlying etiology of AUBs. Individual-level genetic profiles may be useful to point to personalized intervention timelines, particularly for individuals with high alcohol *Problems* genetic risk scores.

## INTRODUCTION

Alcohol use behaviors (AUBs) encompass a wide variety of normative and problematic aspects of alcohol consumption, including typical drinking frequency and quantity, heavy episodic (binge) drinking, intoxication, experience of alcohol-related problems, and clinically significant alcohol use disorders (AUDs). These behaviors are heritable (1–3), with a complex polygenic architecture influenced by many genetic variants primarily of small individual effect (4, 5). Genes exert their effects through both broad influences on individual predispositions towards impulsive, rewarding behaviors (“externalizing”; EXT), as well as processes specific to AUBs such as alcohol metabolism (2, 6–8). By amassing very large samples, genome-wide association studies (GWASs) have begun to identify the genes that influence both broad EXT and alcohol-specific risk (4, 5, 9). However, these studies usually investigate static AUB measures, and it remains mostly unknown how the associated genetic variants interact with developmental processes.

AUBs are dynamic phenomena that unfold in different ways across the lifespan, with strong age-related trends in prevalence as well as substantial between-individual and between-cohort variability in these trends (10–18). Alcohol addiction is similarly dynamic, thought to involve three distinct stages of neurobiological adaptation that correspond to changes in motivations for drinking and compulsivity of alcohol use (19). Genetic epidemiology studies in twins have established that the heritability of AUBs changes across the lifetime, with the relative importance of genes increasing until a plateau in middle adulthood (20), followed by a slight drop-off in older adulthood (21). Some evidence suggests that there are also qualitative changes in which genes affect AUBs at different life stages (15, 22, 23). Certainly, the varying pattern of risk/protective factors for AUBs across the lifespan, such as (legal) access to alcohol, social drinking habits, and prevalence of disorders that co-occur with alcohol use (24), suggests that the underlying causes of AUBs - both genetic and environmental - are developmentally dynamic.

Gene identification studies, however, typically focus on a snapshot of behavior at a single timepoint, such as a composite measure of current consumption quantity, past-year alcohol problems, or lifetime AUD diagnoses (4, 5, 25). This approach, while aiming to maximize statistical power, risks missing out on differences critical to the etiology of AUBs such as longitudinal variation or changes in the relative importance of broad versus specific risk processes across time. Existing evidence indicates that the genetic influences on AUBs are not homogeneous, but can be separated into distinct factors related to consumption, problems, and patterns of AUBs and beverage preferences (26–28), as well as effects that are shared with EXT or are specific to alcohol use (8). EXT-related genes are correlated with early initiation of substance use, while alcohol-specific genes are more relevant for increasing alcohol use/problems over time (8, 29). The precedence of different neurobiological systems at different stages of the addiction cycle (19) also suggests that different sets of genes may be important for initiation, acceleration, and/or persistence of alcohol use. Such different sets of genes have been observed for different stages of smoking behavior and nicotine dependence (4) and are likely to exist for other addictive behaviors.

Understanding the dynamics of how genes and developmental trajectories interact to shape AUBs could point to critical time points in which interventions would be most effective, as well as improving our understanding of the mechanisms driving the statistical associations observed in GWAS. In this study, we leverage longitudinal samples to investigate the dynamic effects of genes on AUBs across the lifespan, from early adolescence to middle and older adulthood. Using polygenic scores (PGSs), we examine how the aggregate genetic influences representing diverse EXT- and AUB-related processes shape trajectories of alcohol use across multiple environmental contexts.

## METHODS

This study was pre-registered under the Open Science Framework (http://osf.io/qg8sa). After registration, two additional cohorts (described below) were added to the analysis plan.

### Participants

Data were obtained from multiple longitudinal studies designed for genetic epidemiology research. Samples include a national study of adolescents in the US (AddHealth), a birth cohort in the UK (the Avon Longitudinal Study of Parents and Children; ALSPAC), a US study of families densely affected with AUDs (the Collaborative Study on the Genetics of Alcoholism; COGA), two twin birth cohorts in Finland (FinnTwin12 and the older Finnish Twin Cohort [FTC]), and an unselected cohort of university students in the US (Spit for Science; S4S). Data collection included DNA samples for genotyping in addition to interviews or self-report surveys of behavior and mental health, collected at repeated intervals. In the current study, we use data from participants with ancestry similar to European reference panels (hereafter referred to as “EUR”) in order to match the genetic ancestry background of the discovery GWASs (9, 26).

#### AddHealth

AddHealth is an ongoing, nationally representative study of adolescents followed into adulthood in the US. In 1994-1995, participants were selected from a stratified sample of 132 schools, resulting in an initial sample of n=90,118 students in grades 7-12. A subset of the original sample (n=20,745) was selected for additional in-home interviews (Wave I/age 11-18 to Wave 5/age 35-42), with a total of n=15,159 individuals providing samples for genotyping at Wave IV. All participants provided informed consent/assent, and the study was approved by the corresponding university Institutional Review Board. Full details on data collection have been previously published (30). The current study includes n=5,107 unrelated EUR participants.

#### ALSPAC

Pregnant women resident in Avon, UK with expected dates of delivery between 1st April 1991 and 31st December 1992 were invited to take part in the study, and the initial number of pregnancies enrolled was 14,541, including 13,988 children who were alive at 1 year of age. After additional recruitment efforts, the total sample size for data collected after the age of seven is 15,447 pregnancies, from which 14,901 children were alive at 1 year of age. Study data were collected and managed using REDCap electronic data capture tools hosted at the University of Bristol (31). The study website contains details of all the data that is available through a fully searchable data dictionary and variable search tool (http://www.bristol.ac.uk/alspac/researchers/our-data/). Ethical approval for the study was obtained from the ALSPAC Ethics and Law Committee and the Local Research Ethics Committees. Consent for biological samples has been collected in accordance with the Human Tissue Act (2004). Full details on data collection have been previously published (32, 33). The current study includes n=5,214 unrelated EUR participants with available genotypic data and measurements collected at ages 16-28.

#### COGA

This study, initiated in 1989, recruited high-risk families through adult probands in treatment for alcohol dependence, including probands, their relatives, and community-ascertained comparison families (n=16,848). A prospective study of a subset of adolescents and young adults (aged 12-22) from COGA families was initiated in 2004, with subjects reassessed every two years. Currently, 89% of participants have 2+ interviews completed. All participants and their families provided informed consent/assent, and the study was approved by the corresponding university Institutional Review Board. Full details on data collection have been previously published (34). The current study includes n=1,955 EUR participants from the prospective sample.

#### FinnTwin12

This population-based study of Finnish twins born 1983–1987 was identified through Finland’s Central Population Registry. A total of 2,705 families (87% of all identified) were enrolled and twins were invited to participate in mailed surveys at ages 12, 14, 17-18, 21-26, and 34-40. An intensively studied subset (1035 families) was selected for genotyping and additional interviews. All participants and their families provided informed consent/assent, including written informed consent for all those who provided blood or saliva samples for DNA genotyping. The study was approved by the Indiana University Institutional Review Board and by the Helsinki University Hospital (HUS) Regional Ethics Committee. Full details on data collection have been previously published (35). The current study includes n=1,219 EUR participants with genetic data passing quality control.

#### FTC

The older FTC is a population-based study of twins born in Finland before 1958. Four waves of surveys were conducted by mail in 1975, 1981, 1990, and 2011. Ethical approval was obtained from the HUS Regional Ethics Committee (ID 01/2011). Informed consent to participate was not obtained directly but was inferred from the completion of the questionnaires. All those who provided blood or saliva samples for DNA have provided written informed consent. The purpose of the study was explained to the participants, and all participants were aware that they could withdraw from the study at any time without any consequences. A full description of the older FTC has been published previously (36). The current study includes n=6,257 EUR participants with genetic data passing quality control and who were aged 18-40 in 1975.

#### S4S

The S4S study recruited incoming students (n=15,067) at a large, urban, public university in the mid-Atlantic US, starting in 2011. All first-time freshmen aged >18 years were eligible to complete an online self-report survey, with follow-up surveys each subsequent spring in which they were enrolled at the university. Data collection was carried out using the secure REDCap system of electronic data capture tools (31). All participants provided informed consent, and the study was approved by the corresponding university Institutional Review Board. Full details on data collection have been previously published (37). The current study includes data from n=4,549 EUR participants from the first 5 cohorts whose DNA has been genotyped and passed quality control procedures.

### Measures

In each cohort (except FTC; see below), we examined measures of typical frequency of alcohol consumption (CON) and heavy episodic drinking (HED), measured in days per month. Each instance of a repeated measure was recorded along with participants’ age at the time of assessment, rounded to the nearest year. As ALSPAC, FinnTwin12, and S4S surveys were administered at structured times, some variability was observed in participants’ age at assessment for each wave. To prevent small cell counts and unreliable model estimates, we generally collapsed these and assigned all individuals to the most common age group within wave (e.g., all S4S participants were coded as age 18 at the first survey wave, while 6% of participants were actually aged 19 and 0.5% older than 19). When sample sizes per age were sufficient, we included multiple age bins (e.g., age ranges of 34-40 in the last wave of FinnTwin12 were recoded as 34-36=35 and 37-40=38). In AddHealth and COGA, we truncated the age range at 32, given the small number of observations outside this range. Based on the low prevalence of drinking initiation prior to adolescence and the ages/waves at which different cohorts had data available, we treated age 14 as the baseline for statistical models and excluded any data collected before this age. A timeline of the assessments for each cohort can be found in **Figure 1** and **Supplementary Table S1**.

**Figure 1.**
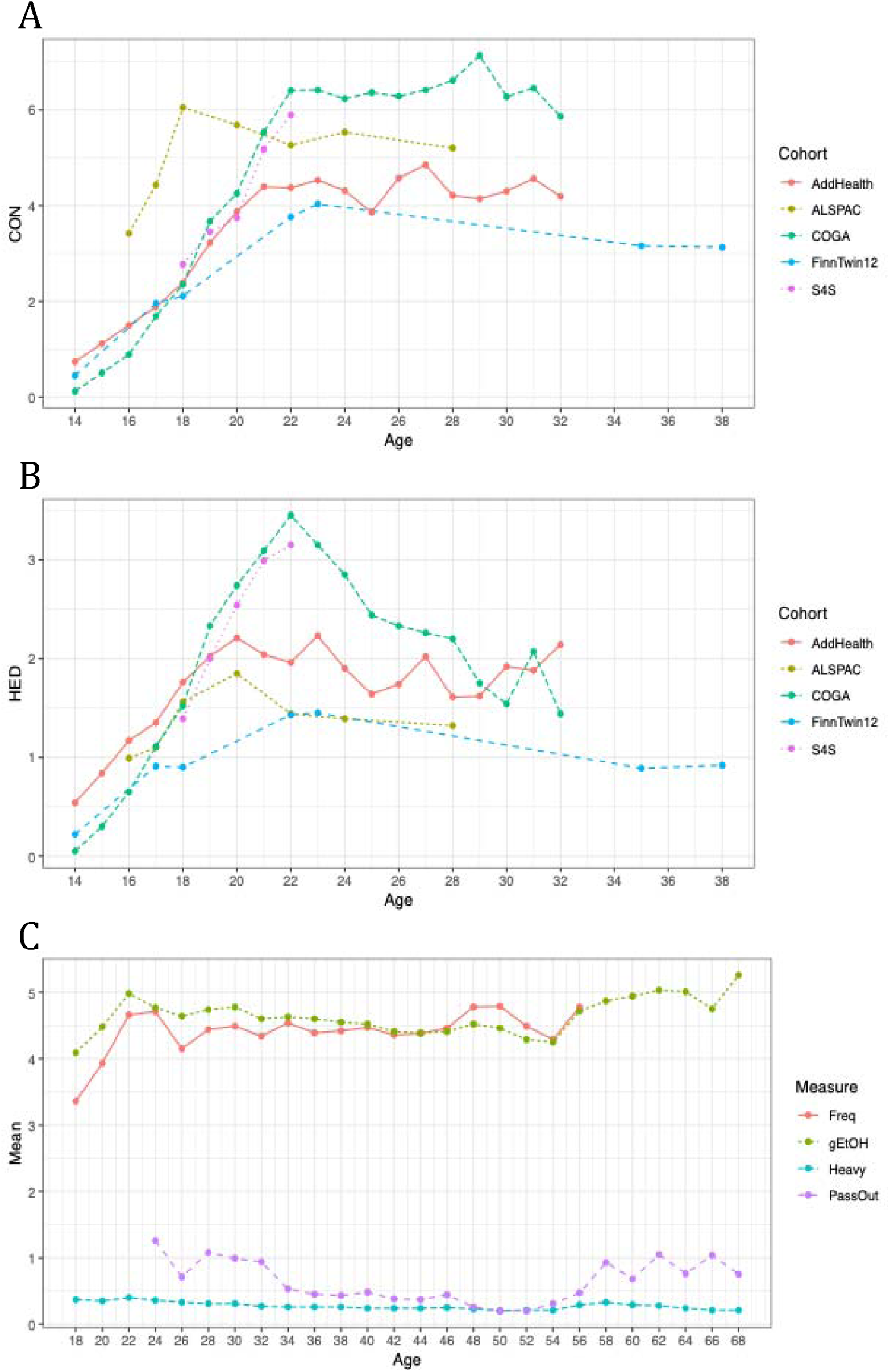
Mean levels per age of A) typical consumption and B) heavy episodic drinking frequencies in five longitudinal cohorts and C) multiple alcohol use behaviors in the older FinnTwin Cohort.

#### CON

Consumption frequency was measured using item #1 of the Alcohol Use Disorders Identification Test (AUDIT; 38) questionnaire (“How often do you have a drink containing alcohol?”) or similarly worded items, recording the number of typical drinking days in categories from “never” to “daily”. These categories were recoded into pseudo-continuous number of days per month, using the median value for categories with a range of days (i.e., 3 days for the category “2-4 times per month”) or the most conservative value when the range was non-specific (i.e., 16 days for the category “4 or more times a week”).

#### HED

Intoxication or HED frequency was measured using item #3 of the AUDIT questionnaire (“How often do you have six or more drinks on one occasion?”) or similarly worded items, recording the number of typical heavy drinking/intoxication days in categories from “never” to “daily”. As for CON, these categories were recoded into pseudo-continuous number of days per month. In FinnTwin12, the HED item was phrased as “How often do you drink so that you get at least slightly intoxicated?” (ages 14 and 17-18), “How often do you drink so that you get really drunk?” (age 21-26), and AUDIT item #3 (age 34-40). In COGA, the HED item was phrased as “How often did you have five or more drinks in 24 hours during the last 12 months?” and recoded to days per month. In S4S, HED was measured in two ways: 1) a sex-specific question “How often do you have [five/four] or more drinks in a single sitting (considered about a 2 hour period)?” for males/females, or 2) the values from the CON measure, above, if participants’ reported typical drinking quantity was >=5 drinks per day (otherwise HED frequency=0). The former measure was only available for a subset of waves; we took the maximum value when both measures were available.

#### Other AUB measures

AUBs were measured somewhat differently in the FTC sample. However, because this is the only cohort with data collected into older adulthood (up to age 67), it provides important additional information about the trajectory of genetic risk later in life. We therefore include data from four AUBs measured in this sample. Frequency of alcohol use (“How often do you drink alcohol?”) was measured for beer, wine and spirits separately and the maximum value across beverage types within wave was taken (Freq) and recoded into a pseudo-continuous count of days per month. gEtOH was a composite measure of grams of ethanol consumption per month, x □ log(x+1) transformed, as previously described (39). Heavy was a dichotomous indicator of whether participants drink “more than five bottles of beer or more than a bottle of wine or more than half a bottle of hard liquor” on the same occasion, at least once a month. Finally, PassOut was a pseudo-continuous measure of the number of days in the past year that participants passed out due to alcohol consumption. Measures were binned in 2-year age intervals to stabilize estimation, and the minimum baseline age (intercept) was 18 for Freq, gEtOH, and Heavy, and 24 for PassOut, since this was assessed in the 1981 and later questionnaires but not the 1975 questionnaire.

#### Polygenic scores (PGSs)

PGSs were derived based on GWAS summary statistics of a prior structural equation model (40), which identified three genetically informative latent factors underlying a set of 18 normative and problematic AUBs. These factors represented 1) chronic and severe alcohol *Problems*, 2) *BeerPref*, a decreasing pattern of alcohol use/problems later in life marked by a preference for drinking beer and drinking without meals, and 3) overall quantity and frequency of *Consumption* of varied alcoholic beverage types. To examine broad as well as alcohol-specific influences, an additional PGS was derived to index 4) *EXT*, based on a GWAS meta-analysis of impulsivity-related traits and substance use (9).

We created these 4 PGSs in each cohort based on previously published GWAS summary statistics. Summary statistics were weighted using PRS-CS (“auto” version) and 1000 Genomes EUR reference panels for estimating linkage disequilibrium, as provided with the software (41). PGSs were calculated in PLINK2 (42) using the ‘--score’ method. Each PGS was entered in a regression model with 10 ancestry principal components and genetic sex as predictors. The PGS residuals, after removing the effects of these covariates, were used for further analyses.

### Data Analysis

All data analyses were carried out using R statistical software (43). First, to examine the overall level of association between the PGSs and AUBs, mean levels of CON and HED were calculated in each sample, averaging across all available waves of data per individual. Each PGS residual was used to predict mean CON and HED in a linear regression model with the lmer() or lm() functions, for cohorts with (COGA, FinnTwin12, FTC) or without (AddHealth, ALSPAC, S4S) relatives, respectively. Regression coefficient estimates were meta-analyzed using random effects inverse variance weighting with the metafor package (44).

The primary analyses involved latent growth curve (LGC) models, structural equation models that use latent growth factors (intercept, slope, quadratic) to represent the mathematical function underlying the covariance between repeated measures. Here, we fit LGCs for each AUB in each cohort using the OpenMx package (45). First, a baseline model containing either intercept+slope (IS) or intercept+slope+quadratic (ISQ) parameters was fit to determine the general shape of the longitudinal trajectories. After comparing the difference in -2*loglikelihood between models with a chi-square test, the best fitting model was selected. Finally, this model was fit sequentially with the addition of each PGS residual as an individual-level predictor of the latent growth factors to determine how genetic influences captured by the PGSs shape AUB trajectories (**Figure 2**). Full information maximum likelihood approaches for model estimation were used to account for missingness and non-normality. A multi-level application of the model (adapted from https://github.com/OpenMx/OpenMx/blob/master/inst/models/nightly/mplus-ex9.12.R) was implemented to account for the non-independence of observations in samples that included relatives by design (COGA, FinnTwin12, FTC). As >90% of participants in each cohort had initiated alcohol use before or during the assessment timeframe, no observations were removed for missing phenotypic values; individuals were coded with a frequency of zero at each non-drinking wave.

**Figure 2.**
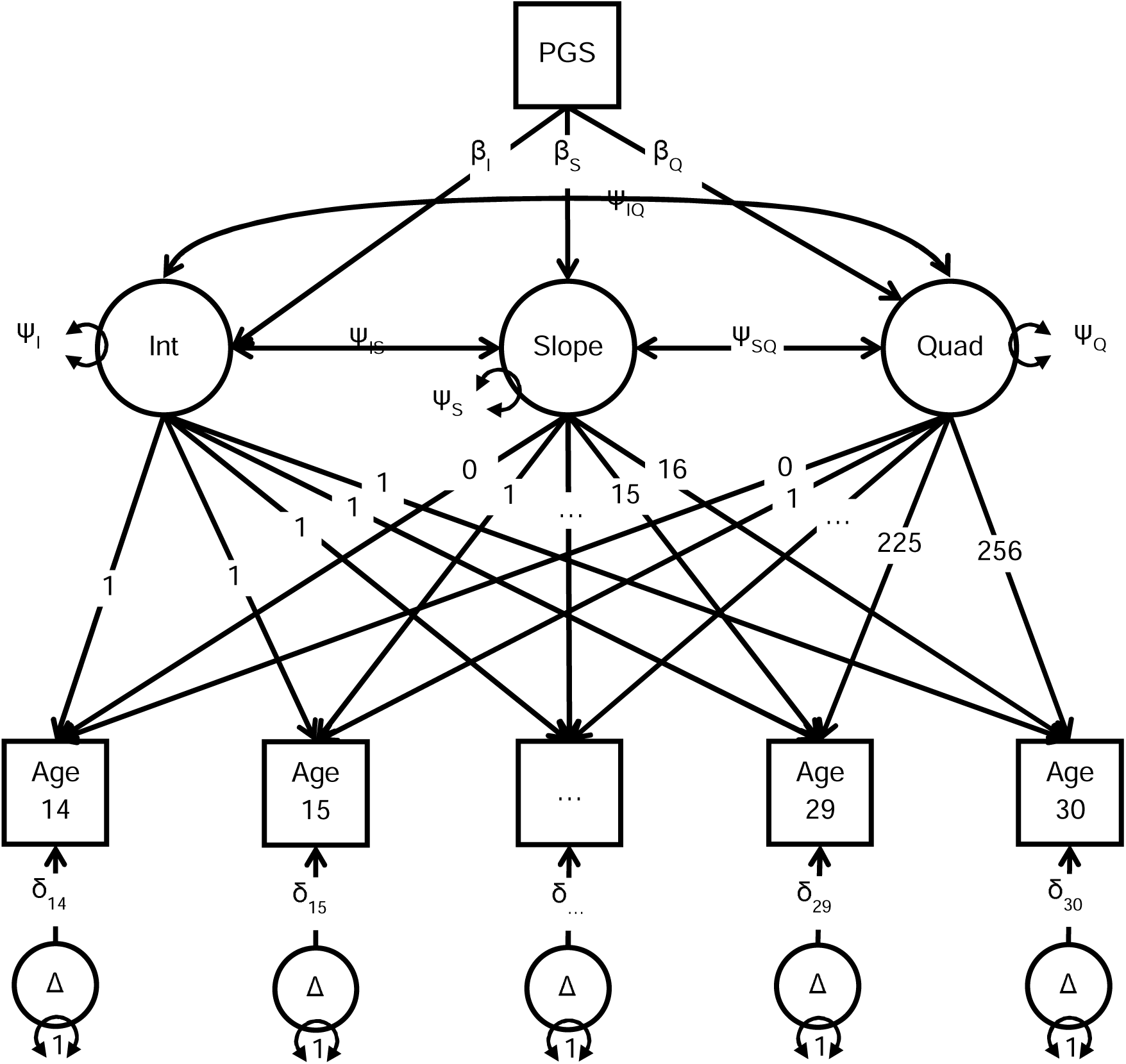
Schematic of the latent growth curve (LGC) model used to estimate the effects of polygenic scores (PGSs) on trajectories of alcohol use behaviors.

Models were tested individually in each cohort and then combined using a random-effects inverse variance-weighted meta-analysis of the PGS effect estimates from each model, using the metafor package. Results from FTC were not meta-analyzed due to the difference in age and available measures, but were compared qualitatively with the primary LGC meta-analysis results. In FTC, only IS models were selected as many participants only had data for 3 or fewer measurement occasions and the quadratic effect estimates were unreliable. We applied a Bonferroni correction for 56 PGS-outcome pairs, resulting in an adjusted alpha=.05/56=.0009.

## RESULTS

Across cohorts, there was a consistent trend of CON and HED increasing sharply from adolescence into emerging adulthood, and then leveling off or decreasing around age 22-24 (**Figure 1a-b; Supplementary Table S1**). This trend was also apparent in the LGC analyses, with the ISQ models, which allow for a quadratic growth factor, providing a better fit than the linear IS models in all cases (p<6.34E-08). Similar longitudinal patterns from age 18+ were observed in the older FTC sample, although there also appeared to be an uptick in AUBs later in life after age 55 (**Figure 1c**).

All PGSs were robustly associated with higher mean levels of CON and HED when these were averaged across time (**Table 1**). The only exception was *BeerPref* PGSs, which were associated with a higher level of HED (meta β=0.064, p=3.65E-05) but not CON. *Consumption* PGSs were the strongest predictor of mean CON (meta β=0.333) while *Problems* and *EXT* PGSs were the strongest predictors of mean HED (meta β=0.154-0.178). In FTC (**Table 2**), *Problems, Consumption,* and *EXT* PGSs were significantly associated with higher mean Freq (β=0.215-0.317) while *Problems, BeerPref,* and *EXT* PGSs were all associated with higher mean levels of gEtOH, Heavy drinking, and PassOut frequency (β=0.025-0.155). Across all analyses, the within-sample variance in AUBs accounted for by each PGS was less than 2%.

**Table 1.**
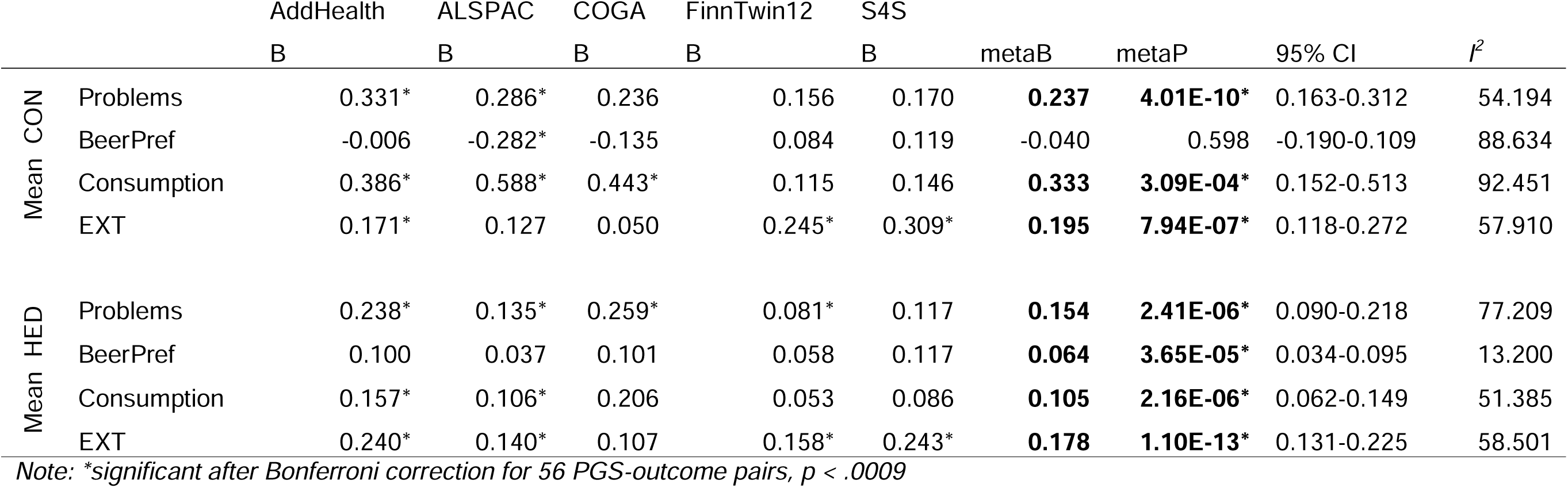
Polygenic score (PGS) prediction of mean levels of typical consumption (CON) and heavy episodic drinking (HED) frequencies, meta-analyzed across five longitudinal cohorts.

**Table 2.** Polygenic score (PGS) prediction of mean levels of alcohol use behaviors in the older Finnish Twin Cohort (FTC).

| | PGS | $\beta$ | P | 95% CI |
| --- | --- | --- | --- | --- |
| Freq | Problems | **0.215** | **3.70E-06\*** | 0.124-0.306 |
|  | BeerPref | 0.118 | 1.14E-02 | 0.027-0.210 |
|  | Consumption | **0.317** | **8.81E-12\*** | 0.226-0.408 |
|  | EXT | **0.250** | **7.72E-08\*** | 0.159-0.342 |
| gEtOH | Problems | **0.089** | **1.03E-04\*** | 0.044-0.134 |
|  | BeerPref | **0.105** | **5.89E-06\*** | 0.060-0.150 |
|  | Consumption | **0.081** | **4.46E-04\*** | 0.036-0.126 |
|  | EXT | **0.155** | **1.65E-11\*** | 0.110-0.201 |
| Heavy | Problems | **0.025** | **4.88E-07\*** | 0.015-0.034 |
|  | BeerPref | **0.034** | **9.54E-12\*** | 0.024-0.043 |
|  | Consumption | 0.016 | 1.03E-03 | 0.007-0.26 |
|  | EXT | **0.046** | **1.01E-20\*** | 0.036-0.056 |
| PassOut | Problems | **0.086** | **4.70E-10\*** | 0.059-0.113 |
|  | BeerPref | **0.069** | **7.50E-07\*** | 0.041-0.096 |
|  | Consumption | 0.016 | 2.57E-01 | -0.012-0.043 |
|  | EXT | **0.084** | **1.10E-09\*** | 0.057-0.112 |

Note: \*significant after Bonferroni correction for 56 PGS-outcome pairs, $p < .0009$

In the LGC meta-analysis (**Table 3**), *Problems* PGSs were significantly associated with a higher slope value for CON (meta β=0.470, p=4.20E-06), meaning that higher genetic risk predicted an accelerated trajectory of increasing drinking frequency over time. There was a general trend that higher PGSs for the genetic dimensions examined were associated with higher intercept and slope values and lower quadratic factor values (i.e., less leveling off of drinking behaviors over time) for both CON and HED, and some were significant in individual cohorts, but these trends were not statistically significant in the combined meta-analysis. Substantial heterogeneity was observed in the estimates across samples, with *I^2^* values of up to 92%. Again, an exception was seen for *Problems* PGSs predicting CON trajectories, which had not only the strongest effects but also the most consistent ones (*I^2^*=0%-2%).

**Table 3.** Polygenic score (PGS) prediction of latent growth factors underlying trajectories of typical consumption (CON) and heavy episodic drinking (HED) frequencies, meta-analyzed across five longitudinal cohorts.

|  | PGS | Factor | AddHealth | ALSPAC | COGA | FinnTwin12 | S4S | Meta-Analysis |  |  |  |
| --- | --- | --- | --- | --- | --- | --- | --- | --- | --- | --- | --- |
| | | | $\beta$ | $\beta$ | $\beta$ | $\beta$ | $\beta$ | Meta $\beta$ | Meta P | 95% CI | i2 |
| CON | Problems | Int | 0.036 | 0.129 | 0.021 | 0.013 | 0.600 | 0.031 | 0.064 | -0.002-0.064 | 0.43 |
|  | Problems | Slope | 0.462 | 0.599 | 0.828 | 0.290 | -2.142 | **0.470** | **4.20E-06\*** | 0.270-0.670 | 2.05 |
|  | Problems | Quad | -0.077 | -0.343 | -0.349 | -0.092 | 2.481 | -0.132 | 0.021 | -0.245--0.020 | 0.02 |
|  | BeerPref | Int | 0.028 | -0.048 | -0.003 | 0.035 | 0.269 | 0.015 | 0.375 | -0.018-0.049 | 0.00 |
|  | BeerPref | Slope | 0.159 | -0.731 | 0.429 | 0.115 | -0.617 | -0.035 | 0.877 | -0.482-0.412 | 74.25 |
|  | BeerPref | Quad | -0.206 | 0.359 | -0.424 | -0.021 | 0.678 | -0.052 | 0.707 | -0.326-0.221 | 70.43 |
|  | Consumption | Int | -0.009 | 0.257* | -0.014 | 0.004 | 0.465 | 0.062 | 0.318 | -0.059-0.183 | 88.55 |
|  | Consumption | Slope | 0.266 | 1.281* | 0.866 | 0.173 | -1.635 | 0.563 | 0.034 | 0.043-1.083 | 79.22 |
|  | Consumption | Quad | 0.257 | -0.709* | -0.142 | -0.053 | 1.874 | -0.105 | 0.596 | -0.494-0.284 | 84.54 |
|  | EXT | Int | 0.146 | 0.287* | 0.063 | 0.028 | 0.587 | 0.127 | 0.022 | 0.018-0.235 | 85.92 |
|  | EXT | Slope | 0.303 | -0.438 | 0.786 | 0.857* | -0.828 | 0.361 | 0.220 | -0.216-0.938 | 83.83 |
|  | EXT | Quad | -0.229 | 0.134 | -0.714* | -0.330* | 0.583 | -0.268 | 0.070 | -0.559-0.022 | 73.74 |
| HED | Problems | Int | 0.055 | 0.090* | 0.025 | 0.007 | 0.432 | 0.042 | 0.025 | 0.005-0.079 | 64.97 |
|  | Problems | Slope | 0.299 | 0.095 | 0.892* | 0.134 | -1.414 | 0.285 | 0.070 | -0.023-0.593 | 82.97 |
|  | Problems | Quad | -0.060 | -0.025 | -0.486* | -0.023 | 1.547 | -0.103 | 0.244 | -0.277-0.071 | 76.52 |
|  | BeerPref | Int | 0.030 | 0.085* | 0.011 | -0.008 | 0.737 | 0.029 | 0.161 | -0.012-0.070 | 70.25 |
|  | BeerPref | Slope | 0.155 | -0.290 | 0.592 | 0.192 | -2.404 | 0.105 | 0.538 | -0.229-0.438 | 85.49 |
|  | BeerPref | Quad | -0.065 | 0.241 | -0.353 | -0.064 | 2.282 | -0.029 | 0.794 | -0.249-0.190 | 85.14 |
|  | Consumption | Int | 0.002 | 0.008 | 0.000 | -0.005 | -0.016 | 0.000 | 0.971 | -0.020-0.019 | 0.00 |
|  | Consumption | Slope | 0.088 | 0.519* | 0.661 | 0.104 | 0.199 | 0.314 | 0.022 | 0.045-0.583 | 77.67 |
|  | Consumption | Quad | 0.118 | -0.374* | -0.333 | -0.023 | -0.003 | -0.140 | 0.236 | -0.373-0.092 | 86.77 |
|  | EXT | Int | 0.141* | 0.174* | 0.010 | 0.024 | 0.576 | 0.091 | 0.029 | 0.009-0.172 | 92.11 |
|  | EXT | Slope | 0.399* | -0.176 | 0.699 | 0.422* | -1.384 | 0.287 | 0.110 | -0.065-0.639 | 87.05 |
|  | EXT | Quad | -0.232 | 0.111 | -0.498* | -0.154* | 1.381 | -0.162 | 0.157 | -0.387-0.062 | 85.99 |

Note: \*significant after Bonferroni correction for 56 PGS-outcome pairs, $p < .0009$

In comparison, LGC models within the older FTC cohort (**Table 4**) demonstrated that *BeerPref* and *EXT* PGSs were associated with a higher intercept for Heavy drinking (β=0.047-0.059, p<1.03E-05). *EXT* PGSs also predicted higher intercept values for gEtOH (β=0.197, p=1.12E-06). These differences likely reflect the older age of the sample, as the intercept now references drinking behaviors in young adulthood, after the period of sharp increase observed in adolescence in other cohorts. Again, no significant prediction from the PGSs was seen for the latent growth factors representing longitudinal changes in AUBs.

**Table 4.**
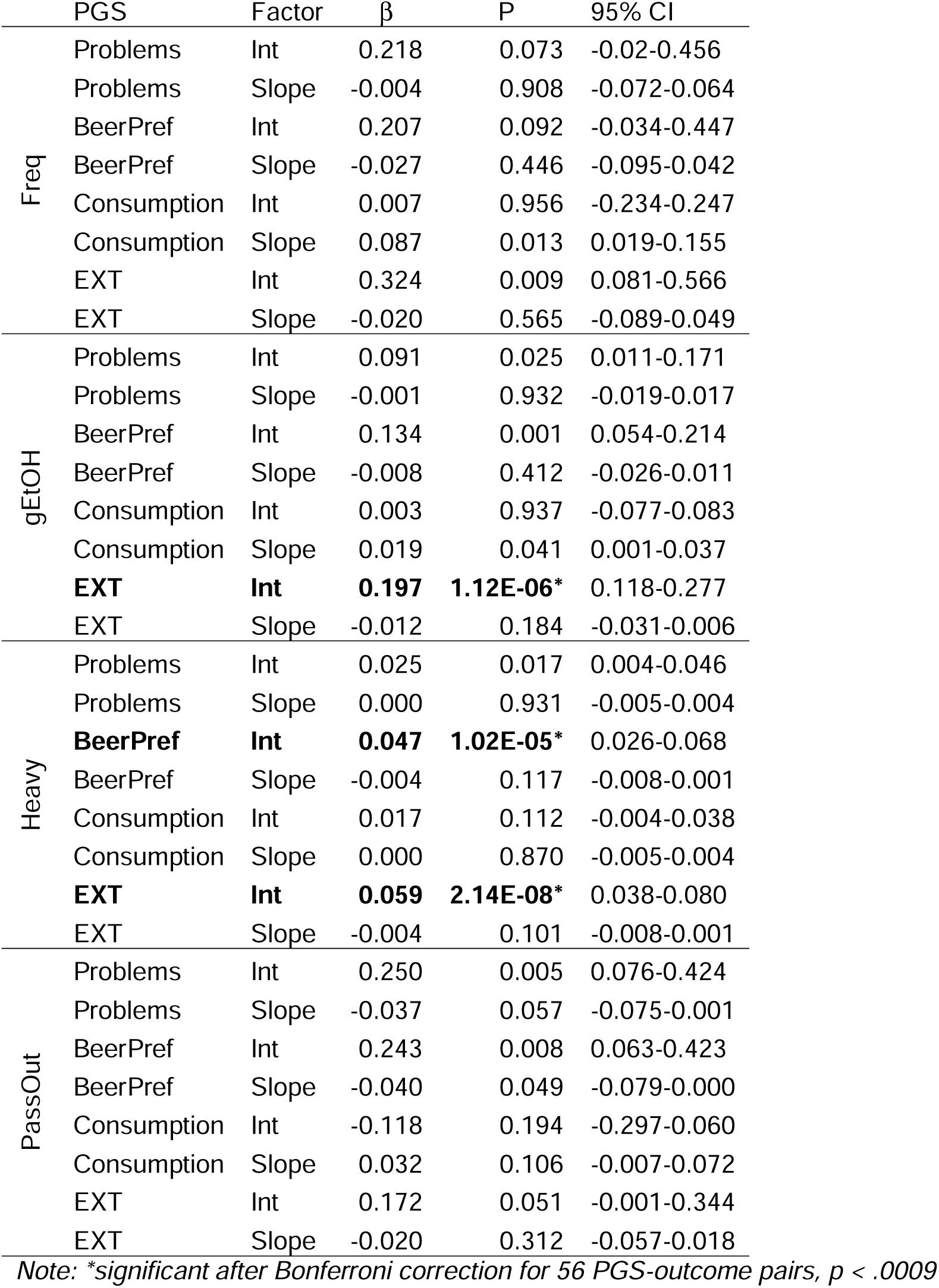
Polygenic score (PGS) prediction of latent growth factors underlying trajectories of alcohol use behaviors in middle and older adulthood in the older Finnish Twin Cohort (FTC).

| | PGS | Factor | $\beta$ | P | 95% CI |
| --- | --- | --- | --- | --- | --- |
| Freq | Problems | Int | 0.218 | 0.073 | -0.02-0.456 |
|  | Problems | Slope | -0.004 | 0.908 | -0.072-0.064 |
|  | BeerPref | Int | 0.207 | 0.092 | -0.034-0.447 |
|  | BeerPref | Slope | -0.027 | 0.446 | -0.095-0.042 |
|  | Consumption | Int | 0.007 | 0.956 | -0.234-0.247 |
|  | Consumption | Slope | 0.087 | 0.013 | 0.019-0.155 |
|  | EXT | Int | 0.324 | 0.009 | 0.081-0.566 |
|  | EXT | Slope | -0.020 | 0.565 | -0.089-0.049 |
| gEtOH | Problems | Int | 0.091 | 0.025 | 0.011-0.171 |
|  | Problems | Slope | -0.001 | 0.932 | -0.019-0.017 |
|  | BeerPref | Int | 0.134 | 0.001 | 0.054-0.214 |
|  | BeerPref | Slope | -0.008 | 0.412 | -0.026-0.011 |
|  | Consumption | Int | 0.003 | 0.937 | -0.077-0.083 |
|  | Consumption | Slope | 0.019 | 0.041 | 0.001-0.037 |
|  | **EXT** | **Int** | **0.197** | **1.12E-06\*** | 0.118-0.277 |
|  | EXT | Slope | -0.012 | 0.184 | -0.031-0.006 |
| Heavy | Problems | Int | 0.025 | 0.017 | 0.004-0.046 |
|  | Problems | Slope | 0.000 | 0.931 | -0.005-0.004 |
|  | **BeerPref** | **Int** | **0.047** | **1.02E-05\*** | 0.026-0.068 |
|  | BeerPref | Slope | -0.004 | 0.117 | -0.008-0.001 |
|  | Consumption | Int | 0.017 | 0.112 | -0.004-0.038 |
|  | Consumption | Slope | 0.000 | 0.870 | -0.005-0.004 |
|  | **EXT** | **Int** | **0.059** | **2.14E-08\*** | 0.038-0.080 |
|  | EXT | Slope | -0.004 | 0.101 | -0.008-0.001 |
| PassOut | Problems | Int | 0.250 | 0.005 | 0.076-0.424 |
|  | Problems | Slope | -0.037 | 0.057 | -0.075-0.001 |
|  | BeerPref | Int | 0.243 | 0.008 | 0.063-0.423 |
|  | BeerPref | Slope | -0.040 | 0.049 | -0.079-0.000 |
|  | Consumption | Int | -0.118 | 0.194 | -0.297-0.060 |
|  | Consumption | Slope | 0.032 | 0.106 | -0.007-0.072 |
|  | EXT | Int | 0.172 | 0.051 | -0.001-0.344 |
|  | EXT | Slope | -0.020 | 0.312 | -0.057-0.018 |

Note: \*significant after Bonferroni correction for 56 PGS-outcome pairs, $p < .0009$

## DISCUSSION

### Summary

Combining multiple international, longitudinal cohorts, we investigated genetic associations with stability and change in alcohol use behaviors across the lifespan. Polygenic scores were used to index multiple AUB-related dimensions, including externalizing behavior (*EXT*), alcohol *Problems*, alcohol *Consumption*, and a preference for drinking beer and drinking without meals (*BeerPref*). Despite their robust associations with AUBs on average, most PGSs were not associated with the dynamics of AUB trajectories. However, *Problems* PGSs significantly predicted a steeper slope of consumption frequency, indicating a genetically influenced acceleration of drinking that is relevant to the development of alcohol problems.

### AUB Trajectories

Consistent with previous longitudinal studies (11, 17, 46), we observed strong age-related trends in prevalence, with AUBs peaking/plateauing in young adulthood. These trends were stable across cohorts separated by continents and decades, despite substantial differences in environments and study designs. Genetic associations with these trajectories, on the other hand, were highly variable, with the meta-analysis *I^2^* statistics indicating that most of the variability in effect sizes for the PGSs was due to between-cohort heterogeneity (47). This highlights an ongoing challenge in utilizing results from GWASs, in which the hundreds of AUB-related genes identified through enormous, pooled samples are still only able to account for a small fraction of the known heritability of AUBs (4, 5), likely due to inter-individual variability in the specific causal genes and processes (45). Despite focusing on AUB dimensions defined with a bottom-up strategy based on their genetic architecture (26), between-sample heterogeneity remained high in our study.

In light of this heterogeneity, it is especially notable that the *Problems* PGSs consistently predicted a steeper slope of consumption frequency across cohorts – the only significant effect in the LGC meta-analysis. This is indicative of a robust biological process driving acceleration of alcohol consumption, and one that may be specific to the peak in emerging adulthood since the same effect was not observed in the older FTC sample. This difference may, however, be a reflection of the lower heritability of AUBs found in older cohorts (18), likely due to secular changes in social constraints on the expression of genetic predispositions. While the genes captured by this latent factor include some in common with other AUBs, such as the well-known alcohol metabolism genes (6), it is only moderately correlated with *Consumption*, *BeerPref*, or *EXT* (26). Further investigation of the genetic influences specific to *Problems* may therefore be helpful to illuminate the causal processes underlying young adult acceleration of alcohol consumption and their links to later alcohol problems.

Results from mean level prediction analyses demonstrated that all four PGSs harbor genetic variants relevant to AUBs, but the growth model results indicated that these may not be the same genes shaping dynamic changes in drinking. Some evidence suggests that, despite the relatively high heritability of AUBs, the genetic influences on trajectories of heavy (episodic) drinking are quite low (17). If so, this may reflect genetic equifinality – high genetic risk leads to high AUB manifestation at some point in life, although the specific timing is shaped by the environment or stochastic effects. Other studies, however, have demonstrated that the heritability of latent growth factors for alcohol frequency and quantity is similar to that of cross-sectional measures of AUBs, ∼30-50% (39, 48). Further research is needed to resolve this question, and it may be necessary to incorporate longitudinal phenotypes into gene discovery efforts to determine whether the same (or any) genes are associated with dynamic versus static AUB measures (49). The continued low variance in mean AUB levels explained by PGSs suggests that other avenues are worth pursuing in an effort to recapture the “missing heritability”, including deep phenotyping (40), longitudinal measures (49), and expanding coverage of the genome to rare and structural variants (50).

### AUB Dimensions

By including multidimensional polygenic scores, we were able to obtain greater insight into the breadth and specificity of genetic effects. PGSs indexing *EXT* and *Problems* were broadly associated with average consumption frequency/quantity as well as heavy (episodic) drinking, while *Consumption* and *BeerPref* PGSs were more specifically associated with average frequency or heavy drinking, respectively. *BeerPref* was even inversely related to consumption frequency in some samples, supporting the hypothesis that this construct represents a genetic predisposition involving a low level of response to alcohol that prompts heavier drinking quantity (40), rather than the social influences that are more relevant to drinking frequency (27). Further, different factors appeared to be most relevant across samples, with, for example, only *Consumption* PGSs showing associations with mean AUBs in the COGA sample but only *EXT* PGSs being associated with mean AUBs in S4S. This heterogeneity across samples further challenges efforts to pinpoint biological mechanisms or improve genetic risk prediction. Our results highlight the need for genetic studies of the diverse heritable risk processes, both broad and specific, that are important for AUBs.

### Conclusions

This study’s findings should be viewed in the context of several limitations, including the lack of sample ancestral diversity, small number of AUB measures which could be harmonized across cohorts/waves, subtle cohort differences in how AUB measures were assessed, and dissection of PGSs which are bound by GWAS discovery power and capture only a small proportion of the phenotypic variance of AUBs. Nonetheless, these are balanced by strengths of the meta-analytic approach drawing from multiple complementary study designs. We highlight the importance of genetic influences on alcohol *Problems* in shaping an accelerating trajectory of drinking frequency, which may already be useful for early identification of at-risk individuals. Multiple other genetic and environmental risk dimensions are, of course, also important in shaping lifelong alcohol use behaviors. Further longitudinal research, especially within gene discovery efforts, is needed to illuminate the mechanisms linking genetic variation to the development of alcohol use behaviors across the lifespan.

## Acknowledgements

This research was funded by a grant to J.E.Savage (VI.VENI.201G-064) from The Netherlands Organization for Scientific Research (NWO). Additional support for data analysis was provided by the U.S. National Institute on Drug Abuse grant T32DA55569 (M.C.) and for each study cohort as described below.

## AddHealth

Add Health is directed by Robert A. Hummer and funded by the National Institute on Aging cooperative agreements U01 AG071448 (Hummer) and U01AG071450 (Aiello and Hummer) at the University of North Carolina at Chapel Hill. Waves I–V data are from the Add Health Program Project, grant P01 HD31921 (Harris) from Eunice Kennedy Shriver National Institute of Child Health and Human Development (NICHD), with cooperative funding from 23 other federal agencies and foundations. Add Health was designed by J. Richard Udry, Peter S. Bearman, and Kathleen Mullan Harris at the University of North Carolina at Chapel Hill.

## ALSPAC

We are extremely grateful to all the families who took part in this study, the midwives for their help in recruiting them, and the whole ALSPAC team, which includes interviewers, computer and laboratory technicians, clerical workers, research scientists, volunteers, managers, receptionists and nurses. The UK Medical Research Council and Wellcome (Grant ref: 217065/Z/19/Z) and the University of Bristol provide core support for ALSPAC. This publication is the work of the authors and Maia Choi and Danielle Dick will serve as guarantors for the contents of this paper. A comprehensive list of grants funding is available on the ALSPAC website (http://www.bristol.ac.uk/alspac/external/documents/grant-acknowledgements.pdf). Genomewide genotyping data was generated by Sample Logistics and Genotyping Facilities at Wellcome Sanger Institute and LabCorp (Laboratory Corporation of America) using support from 23andMe.

## COGA

We continue to be inspired by our memories of Henri Begleiter and Theodore Reich, founding PI and Co-PI of COGA, and also owe a debt of gratitude to other past organizers of COGA, including Ting-Kai Li, P. Michael Conneally, Raymond Crowe, and Wendy Reich, for their critical contributions. This national collaborative study is supported by NIH Grant U10AA008401 from the National Institute on Alcohol Abuse and Alcoholism (NIAAA) and the National Institute on Drug Abuse (NIDA).

## FinnTwin12

FinnTwin12 is supported by the National Institute on Alcohol Abuse and Alcoholism (award numbers R01AA015416, R01AA09203, K02AA018755, and K01AA024152) and the Academy of Finland (grants 100499, 205585, 118555, 141054, 265240, 263278, and 264146).

## Finnish Twin Cohort: older twins

Phenotype and genotype data collection in the twin cohort has been supported by the Wellcome Trust Sanger Institute, the Broad Institute, ENGAGE – European Network for Genetic and Genomic Epidemiology, FP7-HEALTH-F4-2007, grant agreement number 201413., and the Academy of Finland Center of Excellence in Complex Disease Genetics (grant # 352792).

## Spit for Science

Has been supported by Virginia Commonwealth University, P20AA017828, R37AA011408, K02AA018755, P50AA022537, and K01AA024152 from the National Institute on Alcohol Abuse and Alcoholism, UL1RR031990 from the National Center for Research Resources and National Institutes of Health Roadmap for Medical Research, as well as support by the Center for the Study of Tobacco Products at VCU. REDCap support provided by CTSA award UM1TR004360 from the National Center for Advancing Translational Sciences. We would like to thank Dr. Danielle Dick for founding and directing the Spit for Science Registry from 2011-2022, and the Spit for Science participants for making this study a success, as well as the many University faculty, students, and staff who contributed to the design and implementation of the project.

## The Externalizing Consortium

Principal Investigators: Danielle M. Dick, Philipp Koellinger, K. Paige Harden, Abraham A. Palmer. Lead Analysts: Richard Karlsson Linnér, Travis T. Mallard, Peter B. Barr, Sandra Sanchez-Roige. Significant Contributors: Irwin Waldman. The Externalizing Consortium has been supported by the National Institute on Alcohol Abuse and Alcoholism (R01AA015416 – administrative supplement to DMD), and the National Institute on Drug Abuse (R01DA050721 to DMD). Additional funding for investigator effort has been provided by K02AA018755, U10AA008401, P50AA022537 to DMD, as well as a European Research Council Consolidator Grant (647648 EdGe to Koellinger). The Externalizing Consortium would like to thank the following groups for making the research possible: 23andMe, Add Health, Vanderbilt University Medical Center’s BioVU, Collaborative Study on the Genetics of Alcoholism (COGA), the Psychiatric Genomics Consortium’s Substance Use Disorders working group, UK10K Consortium, UK Biobank, and Philadelphia Neurodevelopmental Cohort. The content is solely the responsibility of the authors and does not necessarily represent the views of the funding agencies. The funding agencies had no role in the study design, data analysis, manuscript preparation, or decision to submit for publication.

## Data availability

Raw data from this study are available to qualified researchers. AddHealth: dbGaP accession no. phs001367.v1.p1. ALSPAC: https://www.bristol.ac.uk/alspac/researchers/access/. COGA: https://cogastudy.org/. Finnish Twin Cohort (FinnTwin12 and older cohort): Data used in the analysis is deposited in the Biobank of the Finnish Institute for Health and Welfare (https://thl.fi/en/web/thl-biobank/forresearchers) and is available to researchers after written application and following the relevant Finnish legislation. Spit for Science: dbGaP accession no. phs001754.v4.p2.

